# The impact of COVID-19 on the provision of respectful maternity care: findings from a global survey of health workers

**DOI:** 10.1101/2021.05.05.21256667

**Authors:** Anteneh Asefa, Aline Semaan, Therese Delvaux, Elise Huysmans, Anna Galle, Emma Sacks, Meghan A. Bohren, Alison Morgan, Michelle Sadler, Saraswathi Vedam, Lenka Benova

## Abstract

**Background:** Significant adjustments to the provision of maternity care in response to the COVID-19 pandemic and the direct impacts of COVID-19 can compromise the quality of maternal and newborn care.

**Aim:** To explore how the COVID-19 pandemic affected frontline health workers’ ability to provide respectful maternity care globally.

**Methods:** We conducted a global online survey of health workers to assess the provision of maternal and newborn healthcare during the COVID-19 pandemic. We collected quantitative and qualitative data between July and December 2020 and conducted a qualitative content analysis to explore open-ended responses.

**Findings:** Health workers (n=1,127) from 71 countries participated; and 120 participants from 33 countries provided qualitative data. The COVID-19 pandemic negatively affected the provision of respectful maternity care in multiple ways. Six central themes were identified: less family involvement, reduced emotional and physical support for women, compromised standards of care, increased exposure to medically unjustified caesarean section, and staff overwhelmed by rapidly changing guidelines and enhanced infection prevention measures. Further, respectful care provided to women and newborns with suspected or confirmed COVID-19 infection was severely affected due to health workers’ fear of getting infected and measures taken to minimise COVID-19 transmission.

**Discussion:** Multidimensional and contextually-adapted actions are urgently needed to mitigate the impacts of the COVID-19 pandemic on the provision and continued promotion of respectful maternity care globally in the long-term.

**Conclusions:** The measures taken during the COVID-19 pandemic disrupted the quality of care provided to women during labour and childbirth generally, and respectful maternity care specifically.

## Introduction

Globally, the mistreatment of women during facility-based childbirth is an urgent public health issue that violates women’s rights and contributes to a suboptimal uptake of life-saving maternal health services^1^. Respectful maternity care (RMC), defined by the World Health Organization (WHO) as “the care organised for and provided to all women in a manner that maintains their dignity, privacy and confidentiality, ensures freedom from harm and mistreatment, and enables informed choice and continuous support during labour and childbirth”^2(p3)^ can be a powerful approach to improve the provision of person-centred care, including eliminating the mistreatment of women in health facilities^3^. Promoting RMC also plays a critical role in uptake of care in settings where birthing at home without a skilled attendant is common, due to perceived or actual mistreatment in health facilities^2^. Accordingly, RMC has the potential to reduce global inequities in maternal and neonatal health if promoted from the perspectives of health system strengthening^4^. It is also an essential component of high-quality maternal care^5^, according to the WHO framework for quality care for pregnant women and newborns.

Birth companionship, emotional support, effective communication, pain relief measures (pharmacological and non-pharmacological), respecting women’s preferred birth position, and allowing mobility during labour are recommended practices from the 2018 WHO intrapartum care guideline^2^. These practices form key dimensions of woman-centred care leading to a positive childbirth experience, when complemented by essential resources, including motivated staff, supplies, and infrastructure^6^. RMC is also an essential component of the continuum of care that all women should receive during pregnancy, childbirth, and the postpartum periods. Labour and childbirth are particularly vulnerable times in the continuum and the prevalence of mistreatment is highest in the hours before birth, when the risk of serious complications for both women and babies is highest^7, 8^. Newborns may also experience mistreatment in the immediate postnatal period, which compromises the rights of the newborn and erodes women’s trust in the system and future healthcare utilisation^9^.

Multiple health system barriers, including the shortage of beds, supplies and health workforce, financial bottlenecks, weak referral system, and health workers demotivation jeopardise the promotion of RMC in health facilities^10^. Additionally, shocks such as the COVID-19 pandemic disrupt a health system’s capacity to provide high-quality maternal and newborn health services^11^. An increasing number of reports regarding the impacts of the COVID-19 pandemic on the provision of maternal and newborn care document increased stress, absenteeism, resignation, and redeployment among health workers, affecting the quality of maternity care in health facilities^12, 13^. Additionally, the shift to telemedicine in some settings meant that the care provided to pregnant and breastfeeding women lacked face-to-face interaction, limiting the extent of support provided to women and their newborns; and those living in areas with poor infrastructure were often unable to access care through telehealth technology^14^.

Early on in the pandemic, systemic and structural adjustments were made in the context of limited information about COVID-19 – including on transmission and risk. In many settings, women and their newborns were separated^12^. While interim guidance emerged, including from WHO and Ministries of Health, that all pregnant women regardless of their COVID-19 status should be encouraged to stay with their newborn, commence breastfeeding early and have skin-to-skin contact, some of these harmful practices continued^15^. In many settings globally, women were also prohibited from having a companion of their choice to support them during labour and childbirth, despite the WHO recommendation that all pregnant women—including those with suspected or confirmed COVID-19—have to access companionship^16, 17^. Furthermore, there are reports from several countries that women were subjected to labour induction and caesarean sections without clinical indication during the first months of COVID-19^18^ presumably to reduce women’s length of stay in health facilities, thereby the risk of COVID-19 transmission^19^.

Overall, global health experts across diverse settings warned that adjustments made to the provision of antenatal, intrapartum, and postnatal care as a result of the COVID-19 pandemic and impacts of the COVID-19 themselves could affect women’s and newborns’ rights to respectful care^20^. Notwithstanding numerous COVID-19 research that focussed on infection control and medical complications which could affect perinatal health, reports on the impacts of the pandemic on RMC were only anecdotal. This study is the first global study to examine how the COVID-19 pandemic affected the provision of RMC from the perspective of frontline health workers.

## Materials and methods

To conduct a global study of maternal and newborn health service provision during the COVID-19 pandemic, we designed an online global survey to examine provision of maternal and newborn care services during pregnancy, childbirth, and postpartum periods, seeking perspectives of health workers^21^. Two rounds of online global surveys have been concluded so far—the first round was conducted early in the pandemic (March – July 2020)^12^ and the second round later (July – December 2020); the third-round survey started in December 2020 and will be analysed in 2021. This paper reports on findings from open-ended questions in the second survey round, where specific questions regarding RMC were included.

### Study design and recruitment

This study is part of a broader multilingual survey that used a repeated cross-sectional design. Data were collected using an online self-administered questionnaire prepared using the KoBoToolbox toolkit. The survey was advertised globally using different communication methods (WhatsApp, Twitter, newsletters) and networks (national and international societies, maternal and newborn study consortia members, etc.) to ensure a broad representation of maternal and newborn health workers.

### Questionnaire development

The questionnaire was developed by a multi-disciplinary team of international experts in maternal, sexual and reproductive health, health systems, epidemiology, sociology, anthropology, and clinical practice. After analysis of the first round responses, the study team adapted the questionnaire in light of responses that suggested essential components of RMC were negatively affected, and emerging information from the dynamic pandemic situation. A revised questionnaire was used in the second round. The core structure of the survey was maintained with some modifications and additions of RMC focused questions to explore this theme that emerged in the responses received during the first round and other issues relevant to the pandemic at that time. The main RMC-focused question that was added in the second survey was *‘At this time, to what extent do you feel that you are able to provide respectful care to women and newborns compared to before the COVID-19 outbreak?’* (5-point Likert scale). The questionnaire was prepared in English, and translated into 11 languages (Arabic, Chinese, Dutch, French, German, Italian, Japanese, Kiswahili, Portuguese, Russian, and Spanish) by experts fluent both in English and the language into which they translated the questionnaire. The full version questionnaire is available on the study website^21^.

### Participants and procedures

Health workers, mainly midwives, obstetricians, gynaecologists, nurses, and general practitioners, who self-identified as providing maternal and newborn care services during the COVID-19 pandemic across the globe were eligible for inclusion in the study. From the total survey participants, those who responded to the main RMC question were included in this current analysis. Participants who reported that their ability to provide RMC was about the same, somewhat better, and substantially better were not asked any further questions regarding RMC. On the other hand, participants who responded that their ability to provide RMC during the pandemic was somewhat lower or substantially lower were asked an open-ended question to explore how the pandemic affected their ability to provide RMC; these responses are included in this analysis. We also reviewed responses of all participants to other open-ended questions in the questionnaire and identified those that referred to experiences related to RMC during the pandemic, and included these responses in the qualitative analysis.

### Data management and analysis

Data were exported from the online server to Microsoft Excel and cleaned by the first author. Participant responses with more than 90% missing across survey items were excluded. Non-English text responses were then translated to the English language by AS (French and Arabic), bilingual members of the larger study working group (Japanese, Italian, and Russian), and AG (Portuguese and Spanish). Qualitative data were analysed using content analysis technique adhering to the following four recommended steps^22, 23^. First, reading and re-reading of the open-ended responses was done by AA to understand the data and generate preliminary coding. Second, the codes were reviewed by ES vis-à-vis the responses to improve the reliability of the methods used^23^. Third, the agreed-upon codes were organised by AA to identify preliminary themes and codes. Fourth, the themes were reviewed by the research team to check for congruence with the data; two themes were merged as they had substantial overlapping contents^23^. In reviewing the themes, we selected codes related to respectful nature of the care provided to women and/or newborns with suspected or confirmed COVID-19. We report components of the study using the standards for reporting qualitative research (SRQR) to ensure rigour and credibility ^24^.

## Findings

From the total 1,248 maternal and newborn health workers who participated in the second survey round, 1,127 (90.3%) participants from 71 countries responded to the respectful care question (Table 1). Among these, 192 (17%) reported that their ability to provide respectful care during the COVID-19 pandemic was somewhat lower or substantially lower than before the pandemic; the remaining participants reported that their ability to provide respectful care was about the same (29.6%) and somewhat better or substantially better (46.8%). Perceived somewhat or substantially lower ability to provide respectful care during the pandemic varied across participants’ countries income groups; it was highest among those who worked in high-income countries (24%), followed by those from low-income countries (21%), and middle-income countries (14%). somewhat or substantially lower ability to provide respectful care was also higher among midwives (24%) compared to obstetricians and gynaecologists (15%), and nurses (10%). Among the 192 respondents who reported their ability to provide respectful care was somewhat or substantially lower, we analysed the responses of those who completed more details in the open-text follow-up question (n=120, Supplementary file 1). Of these 120 participants, 52% were from high-income countries, 37% from middle-and 12% from low-income countries (Table 1).

**Table 1.** Characteristics of sample of respondents whose experiences were analysed in this paper.

| Characteristics | (A) Full sample of respondents<br>(n = 1127), n(%) | (B) Subset of A:<br>those with lower or<br>substantially lower<br>ability to provide<br>respectful care<br>(n = 192), n(%) | (C) Subset of B:<br>those provided<br>responses to the<br>open text question<br>(n = 120), n(%) |
| --- | --- | --- | --- |
| Profession |  |  |  |
| Midwife / nurse-midwife | 280 (24.8) | 68 (35.4) | 46 (38.3) |
| Obstetrician/gynaecologist | 242 (21.5) | 37 (19.3) | 30 (25.0) |
| Other medical doctor | 219 (19.4) | 37 (19.3) | 24 (20.0) |
| Nurse | 316 (28.0) | 33 (17.2) | 13 (10.8) |
| Other/no response | 70 (6.2) | 17 (8.9) | 7 (5.8) |
| Gender |  |  |  |
| Female | 873 (78.4) | 148 (77.1) | 87 (71.9) |
| Male | 233 (20.9) | 42 (21.9) | 31 (25.8) |
| Other/prefer not to say | 8 (0.7) | 2 (1.0) | 2 (1.7) |
| Participant's country income status |  |  |  |
| High-income | 305 (27.1) | 73 (38.0) | 62 (51.7) |
| Middle-income | 711 (63.1) | 96 (50.0) | 40 (33.3) |
| Low-income | 111 (9.8) | 23 (12.0) | 18 (15.0) |
| Position |  |  |  |
| Team member or other | 709 (65.5) | 117 (62.6) | 66 (55.5) |
| Head of team or department | 209 (19.3) | 38 (20.3) | 29 (24.4) |
| Independent or privately-practicing | 120 (11.1) | 26 (13.9) | 22 (18.5) |
| Head of facility | 44 (4.1) | 6 (3.2) | 2 (1.7) |
| Participant's facility provided care to<br>women or newborns with COVID-19<br>suspected or confirmed cases since the<br>beginning of the pandemic |  |  |  |
| No | 280 (25.3) | 62 (33.0) | 45 (37.5) |
| Yes, to both confirmed and<br>suspected cases | 266 (24.0) | 41 (21.8) | 34 (28.3) |
| Yes, to confirmed cases | 268 (24.2) | 35 (18.6) | 18 (15.0) |
| Yes, to suspected cases | 184 (16.6) | 30 (16.0) | 14 (11.7) |
| Do not know | 109 (9.6) | 20 (10.6) | 8 (6.7) |

Our qualitative analysis identified the following six themes of how the pandemic negatively affected health care workers’ ability to provide RMC: less family involvement, reduced emotional support to women, reduced physical support for women, compromised standards of care, increased risk of medically unjustified caesarean section, and overwhelmed staff with rapidly changing guidelines and enhanced infection prevention measures. The last theme is overarching—it affects the remaining themes and influences the way health workers provide care to women and newborns (Figure 1).

**Figure 1.**
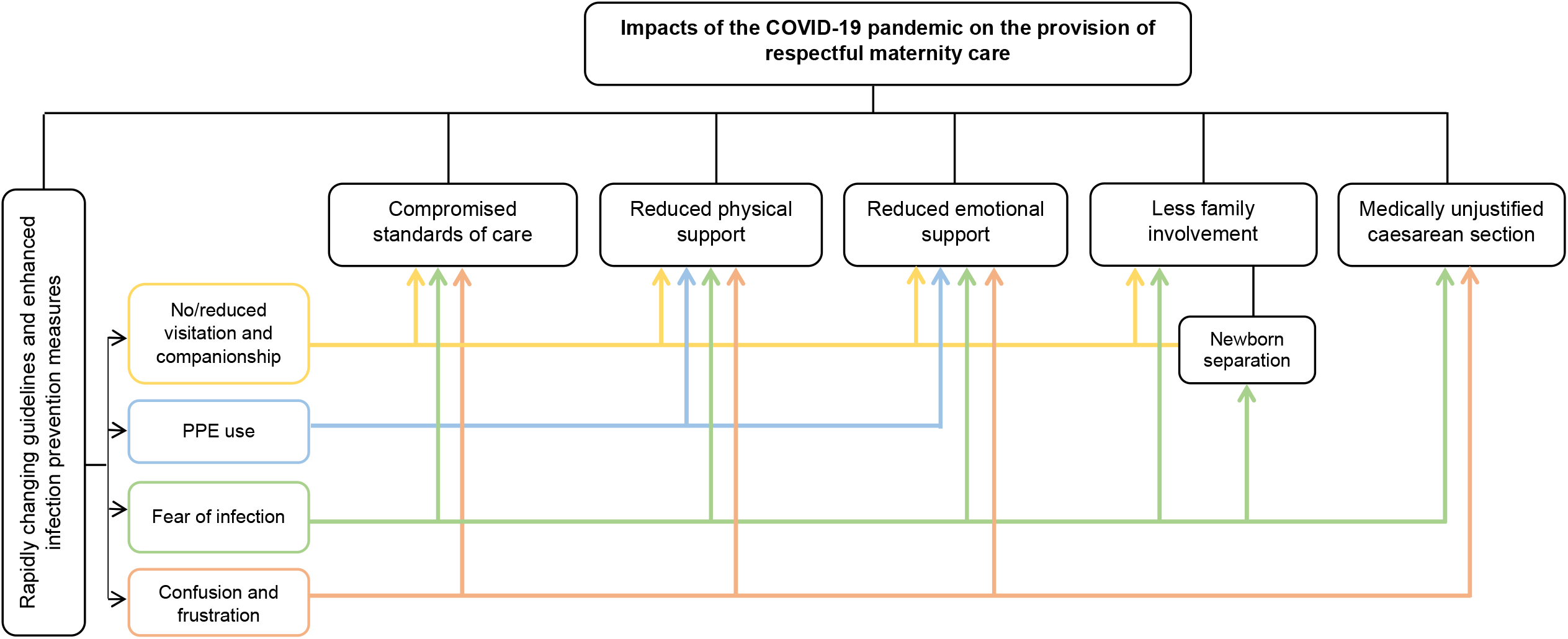
Themes and connections between themes showing impacts of the COVID-19 pandemic on the provision of respectful maternity care globally.

## Less family involvement

Twenty-six participants (among which ten were midwives and nine were obstetricians and/or gynaecologists) from 11 countries (Belgium, Canada, France, India, Italy, Japan, Nigeria, Norway, Spain, Sweden, and United States) discussed issues belonging to this theme.

### Denial of birth companionship

Sixteen participants from various countries reported that enhanced infection prevention measures introduced as a result of the COVID-19 pandemic resulted in the prohibition of the presence of birth companions during labour and childbirth. In settings where partners and/or doulas are typically present for birth, such as Japan or Canada, rules changed to suspend this practice, creating distress among women and their families. To compensate for this, some health workers made phone calls to try to reassure parents who could not witness the birth of their child. In settings where it is customary for women’s mothers to provide care and support during labour and childbirth, the imposed restriction prevented women from benefiting from their mothers’ companionship.

*“The main challenge continues to be the limitation imposed on visiting hours and on companions presence during labour and childbirth*.*”* (Midwife, district hospital, Spain)

Women who underwent caesarean section were also highly disadvantaged due to the restriction to admit companions as they required additional support to recover from surgery during their inpatient stay, and lacked mobility to take care of their newborn, including breastfeeding and early skin-to-skin contact.

*“Accompaniment in caesarean sections and skin-to-skin care after caesarean sections were affected as partners were not allowed to accompany women and there were limited staff to support women*.*”* (Obstetrician/gynaecologist, referral hospital, Uruguay)

In some settings where companions were allowed intermittently depending on facility circumstances, participants reported that shortage of supplies required to undertake frequent cleaning and disinfection and lack of space in maternity wards made it difficult to allow companions. The restriction on companionship was also heightened in scenarios where there was a COVID-19 suspected woman or newborn present in the health facility, due to the priority to prevent COVID-19 transmission. In settings where women are in larger, shared spaces on a prenatal ward, some women were sent back to their homes when there was a COVID-19 suspected or confirmed person present, let alone allowing women to have a birth companion.

### Restrictions on postnatal visitation

In addition to the ban on companionship in labour wards, visiting hours for postnatal wards were substantially reduced and that led to women being left alone and without support in health facilities. Women who required additional pain relief measures, for example those who had lactational mastitis, were severely affected due to the absence of partners, family members or relatives who would have provided supportive care.

*“The partner is not allowed to be with the woman before and after birth on visits*.*”* (Midwife, birth centre, Sweden)

Even with the presence of a companion—in settings where women were allowed to have one—women with complications who required additional support were affected as their relatives or friends, who would have provided periodic support, were not allowed to enter facilities due to the cancelation of visiting hours.

*“…visits as before are not allowed (women can have only one person during the stay)*.*”* (Midwife, clinic, France)

### Newborn separation from parents or family members

In addition to excluding birth companions, ten participants reported that newborns were also kept in separate rooms and women’s partners and family members had reduced access to the newborns. This was reported as a universal practice, not only for women with suspected or confirmed COVID-19. Accordingly, the ban on companions and visitors affected not only the support provided to women during labour and childbirth but also the parental support that would have been provided to newborns during the early stages of life.

*“Some women spend most of their labour without a supportive partner. The infant’s first days of life also passed without the presence and support of the partner*.*”* (Midwife, referral hospital, Italy)

The separation of newborns from their families also affected the collaborative effort between health workers and other family members that health workers would have enjoyed. A nurse from Norway said “*Only one parent is allowed to follow the newborn to the newborn unit. I miss the cooperation with both the mother and father*.”

### Reduced emotional support to women

Nineteen participants (among whom 15 were midwives) from 11 countries (Belgium, Brazil, Canada, France, Germany, Italy, Nigeria, Spain, Switzerland, United Kingdom, and United States) reported issues related to reduced emotional support for women. Emotional support was compromised through three distinct pathways: women not being allowed birth companions with them during labour and childbirth; the ban on cultural mediators such as translators or doulas to support women of culturally and linguistically diverse backgrounds; and health workers themselves limiting personal contact with women in order to reduce infection transmission, largely due to not having sufficient personal protective equipment (PPE). However, even when sufficient PPE was available, health workers reported that providing care and support while wearing face masks and shields created a significant communication barrier.

*“I have the impression that it is more difficult to reassure the patient; the mask creates distance and distrust in the relationship*.*”* (Midwife, clinic, France)

Participants reported that new clinical guidelines introduced in response to the pandemic required health workers to minimise the duration of contact with women, which severely affected the amount and quality of emotional support provided. The new measures also included the prohibition of partners or relatives as birth companions in health facility settings which will be reported in detail later in this paper.

*“Due to the need to maintain physical distance during conversation, examinations are carried out very quickly in order to quickly restore the distance, with no body contacts, such as holding hands, touching arms, comforting etc*.*”* (Midwife, privately-practicing, Germany)

Women who did not speak the official language of the country in which they gave birth were disproportionately affected as translators were not allowed to be present. Instead, when possible, these translators functioned from distance through telephone assistance.

In settings where the supply of PPE was suboptimal, health workers reported having reduced face-to-face contact with women compared with the pre-pandemic level, altering the nature of emotional support and reassurance they would have liked to provide to women. This was further intensified because of health workers who themselves were overwhelmed with multiple responsibilities and emotionally drained as a result of the pandemic response.

*“With patients; we no longer spend time physically. We limit the face-to-face contact. That emotional comfort is not there anymore. Physically examination is no longer practiced as before*.*”* (Obstetrician/gynaecologist, referral hospital, Nigeria)

### Reduced physical support for women

Sixteen participants (among whom nine were midwives) from 11 countries (Belgium, Brazil, Canada, France, Germany, Italy, Nigeria, Norway, Spain, Switzerland, and Unites States, and Uruguay) shared experiences related to this theme. Respondents noted that the pandemic affected the physical support they were able to provide to women during labour and childbirth in different ways. First, the practice of continuous support to women in labour was substantially affected by physical distancing measures. Some women in early labour who would have been admitted and received reassurance, comfort measures and support were sent back home. Second, the COVID-19 pandemic led to staff shortages, thus overwhelming health workers on duty. Further to these, due to the challenges of social distancing and increased requirements for cleaning, women in labour were required to stay in the bed or room to which they were admitted, and not move around or transfer to larger rooms to utilise facilities such as bathtubs during their labour. In some settings, rooms designed for these utilities were repurposed for other uses due to space limitation as a result of the pandemic.

*“The availability of the spaces during labour is much more complicated and cleaning is difficult; so, mothers stay where they have arrived, without being able to go to other rooms where there is a bathtub for example*.*”* (Midwife, district hospital, Spain)

In some settings, women were required to check with health facilities for bed availability before arriving at these facilities. There were instances where labouring women arrived at hospitals where beds were fully occupied and women were left in waiting areas or sent back to their home or to search care at other hospitals. As a result, women could not receive timely physical support from health workers prior to birth, and instead remained without care at their home, on their way to health facilities and even in waiting areas in health facilities.

*“…we have inadequate space to keep pregnant women who have confirmed COVID case and other pregnant women are being sent to their home who come to await for delivery at our facility because of this pandemic; as a result, their care is being compromised*.*”* (Midwife, district hospital, Malawi)

At times when there were confirmed or suspected COVID-19 cases in maternity units, the staffing shortage resulted in compromised care for other women during labour and childbirth—health workers assisting these confirmed and suspected cases could not move to different rooms and assist other women; instead, available staff did whatever they could despite being overworked and fatigued.

*“… in addition, the workload represented by taking charge of a woman in isolation means that we are much less available for other women, the work of other colleagues is also heavily impacted: assistance to the midwife in isolation, taking charge of more patients because the midwife in isolation cannot take on as many follow-ups as usual*.*”* (Midwife, facility type not provided, Belgium)

### Compromised standards of care

Sixteen participants (among which eight were midwives and five were obstetricians and/or gynaecologists) from 13 countries (Argentina, Democratic Republic of the Congo, France, Germany, India, Italy, Japan, Malawi, Nigeria, Norway, Panama, United States, and Uruguay) reported issues belonging to this theme. Participants reported that they gave their maximum effort to provide RMC during the COVID-19 pandemic. Nonetheless, the quality of preventive and counselling services such as breastfeeding counselling and newborn care and support were reportedly declined. Care for newborns, especially those who required additional support, was also affected as their stay in hospital was significantly reduced from what is recommended and they were kept in separate rooms from their mothers in some settings. Ten respondents also reported that breastfeeding support sessions were cancelled.

*“…due to the pandemic, we are not able to practice kangaroo mother care and mother-infant bonding for low-birth-weight babies*.*”* (Neonatologist, referral hospital, Argentina)

*“Although we know that breastfeeding and support must be maintained, due to logistics in my hospital, it is not being carried out and mothers are being separated from their babies*.*”* (Obstetrician/gynaecologist, privately-practicing, Panama)

Health workers reported that vaginal examinations to assess cervical dilation and manual foetal heartbeat monitoring in settings where dopplers were absent were conducted in longer intervals than recommended (every four hours for vaginal examination). Additionally, women and their newborns were being discharged earlier than usual to reduce the chance of COVID-19 transmission in health facilities.

*“Monitoring of foetal heartbeat has reduced because we do not have adequate foetal dopplers and people are afraid of using fetoscopes because of fear of getting infected. Postnatal stay has also reduced from 48 hours waiting to 12 hours for those having a normal spontaneous vaginal birth*.*”* (Midwife, privately-practicing, Malawi)

Consequently, the care women and their newborns received in the immediate postnatal period including breastfeeding education and counselling, mother-infant attachment support, and infant care and support education sessions for fathers was compromised. These inconsistencies and reduction in standards of care were reportedly partly due to the lack of clear and contextualised guidelines on how to provide maternal and newborn care in the face of the COVID-19 pandemic.

*“I feel we are unable to offer good advice to patients as there is no cohesive policy or guideline about how we are addressing the risks of COVID-19 to pregnant and postpartum women*.*”* (Midwife, referral hospital, United States of America)

Women with suspected or confirmed SARS-CoV-2 infection were reported to be the most likely to receive suboptimal care, as the fear of getting infected among health workers and the need for minimising contact was heightened. Moreover, the extra time required for donning and doffing of PPE while caring for these women resulted in depersonalisation of care and limited free movement of health workers leading to low quality of interpersonal and clinical care. The need to use protective devices such as face masks for long hours also created additional burden and discomfort among health workers further affecting their ability and morale to provide RMC.

*“I feel that pregnant women who are suspected of having close contact have many restrictions from normal medical care, such as changes in delivery methods*.*” (Obstetrician/gynaecologist, referral hospital, Japan)*

*“…preventive measures while caring for COVID-19 cases reduced the visibility and movement of service providers due to the protective devices used*.*”* (Midwife, privately-practicing, Italy)

*“All patients with confirmed COVID-19 are being discriminated, no one wants to help them in fear of getting the disease since we have no proper PPE*.*”* (Midwife, district hospital, Malawi)

Similar to the care for women, the care provided to newborns was affected in several ways. Newborns of women with suspected or confirmed COVID-19 had less frequent follow-ups and supportive care. Additionally, recommendations to initiate early skin-to-skin contact to facilitate breastfeeding and attachment were less likely to be followed during the pandemic. Participants also reported that newborns admitted in intensive care units, especially in high-volume hospitals, were likely to be left without attention due to the impact the pandemic had on staff availability.

*“It is challenging to provide care for newborns with COVID-19 or who have contact with COVID-19 positive mothers who need to be in the newborn unit where they will be isolated since there is not any newborn unit set for them”* (Medical Doctor, referral hospital, Kenya)

*“We find some babies are left unattended, especially if they are not as ‘robust’ to be noticed by loud cry…. NAS (neonatal abstinence syndrome) babies are always getting attention and quiet babies have been found to be not touched, held or interacted by adults as often*.*”* (Advanced neonatal nurse practitioner, referral hospital, United States of America)

### Increased risk of medically unjustified caesarean sections

Nine participants from six countries (Argentina, Kenya, India, Japan, Nigeria, and United States) mentioned issues relevant to this theme; two were midwives and four were obstetricians and/or gynaecologists.

Respondents reported that options given to women to choose their preferred birthing position, comfort measures, and mode of childbirth were affected. Participants from several countries (Kenya, Nigeria, Cameroon, United States of America, India, and Argentina) reported that there were higher than usual rates of caesarean sections in their health facilities. The pandemic affected the decision pathway to caesarean section in two aspects. In high-volume maternity care settings where health workers were expected to frequently change PPE due to attending numerous patients, the speed to make decisions to undertake caesarean sections was increased. Routine protocols to assure pre-surgery, interprofessional discussions about options and rationale for caesareans were suspended.

*“…more focus on infection prevention which results in less face-to-face time with COVID-positive mothers; lower thresholds for caesarean sections in COVID-positive mothers which lead to more unnecessary caesarean sections; lack of understanding of principles of disease transmission among both patients and providers/nursing staff*” (Midwife, referral hospital, United States of America)

On the other hand, in lower volume maternity care settings where the consumption of PPE for caesarean sections was higher compared with vaginal births, respondents noted that caesarean sections were delayed despite the presence of indications, endangering women’s lives. Additionally, caesarean sections for women with a COVID-19 positive test could not be performed timely as a separate operating theatre was not available or it was difficult to set one up due to resource limitations.

*“…there is decreased elective inductions and elective caesarean sections, only one labour companion permitted per patient, and prohibition of the use of nitrous oxide for labouring women (this has now been amended, and only patients who are COVID-positive are not permitted to use nitrous oxide)”* (Midwife, referral hospital, United States of America)

*“I have big concern for where pregnant women with COVID-19 in need of caesarean section will deliver because there is no separate theatre for them”* (Medical Doctor, referral hospital, Kenya)

### Providers overwhelmed by rapidly changing guidelines and enhanced infection prevention measures

Six participants from five countries (Democratic Republic of the Congo, Germany, Spain, United Kingdom, and United States) reported issues relevant to this theme; five were midwives.

The COVID-19 pandemic drained health workers physically and emotionally as they were required to cope with multiple and changing guidelines that were introduced to prevent infections in health facilities and provide care for COVID-19 suspected and infected women and newborns.

*“We have less information regarding the protocols, which are still changing. Different protocols in different areas: for example, in the emergency room if clients do not have COVID-19 symptoms, service providers do not use a gown or screen, in the maternity ward they do*.*”* (Midwife, district hospital, Spain)

A midwife from Germany also said that there are “*far too many different recommendations and commands*” with which health workers struggled to cope and implement. Furthermore, social distancing in health facilities and the use of PPE made communication among health workers and between health workers and women tedious and affected the RMC provided to women.

*“The use of personal protective equipment and social distancing is a barrier to communication and makes care difficult and uncomfortable*.*” (Midwife, privately-practicing, United Kingdom)*

A midwife from Italy reported that they were at times confused about how to take care of COVID-19 suspected women and newborns as there was “*lack of a clear pathway to handle COVID-19 cases”* in health facilities. Furthermore, unclear guidelines and uncertainties around the potential consequences of the COVID-19 virus on the health of women and newborns also led to doubt among health workers with respect to prioritising those who need urgent care first.

*“General uncertainty due to the unknown virus leads to uncertainty on all sides, and that has shifted priorities in care*.*”* (Midwife, referral hospital, Germany)

Accordingly, uncertainties and confusion that emerged as a result of the COVID-19 pandemic had the capacity to affect the equitable treatment of women and newborns in health facilities as health workers’ prioritization of clients was not based on robust guidelines. The introduced guidelines and other health system limitations that the participants reported severely affected the ability to provide respectful and supportive care to women and newborns with suspected or confirmed COVID-19 and disadvantaged groups (minors, women who did not speak the official language of the country they gave birth in, and women with developmental or intellectual disabilities).

## Discussion

Documenting evidence of how the COVID-19 pandemic affected the provision of RMC is important not only to prepare for future health system shocks but also to develop evidence-based strategies to augment the progress towards the Sustainable Development Goals. This is the first global study of maternal and newborn health workers to examine the most important ways in which RMC has been affected. The survey allowed health workers to share their experiences in their own words; showing that the effects of the COVID-19 pandemic were multidimensional, negatively reinforcing, and affected both interpersonal and clinical aspects of care to women and their newborns. All six themes align with reductions in essential components of high-quality care as described by the WHO standards for quality of maternal and newborn care^5^. Additionally, the themes extend across four of Bohren and colleagues’ seven domains of mistreatment of women during facility-based childbirth: stigma and discrimination, failure to meet professional standards of care, poor rapport between women and providers, and health system conditions and constraints^1^. Most of the impacts were a result of both suboptimal pre-existing health system and facility-level conditions as well as poor pandemic-related decisions in regard to continued provision of maternity care. They also directly and deeply affected the health workers.

Our study revealed that simple but lifesaving interventions (early breastfeeding, birth companionship, and kangaroo mother care) could not be ensured during the COVID-19 pandemic due to the introduction of restrictive guidelines. These findings are in line with a global survey that examined healthcare providers’ experiences of newborn care provision during the COVID-19 pandemic^27^. We believe that these and some of the affected critical components of RMC during the COVID-19 pandemic were those that could be preserved with little effort and minimal risk, at least after the initial shock of the pandemic. For example, promoting early breastfeeding and ensuring access to a birth companion for all women regardless of their COVID-19 status are strongly recommended practices by the WHO as their benefits outweigh the risks^16^.

During the early COVID-19 pandemic, there were reports of increased caesarean sections on mother’s request to enable partners’ presence at birth in the United Kingdom because of restrictions on the amount of time partners can stay during normal birth^25^. That later led to a blanket ban on “maternal request caesarean sections” and debates on the need to balance between clinical justifications and women’s rights and circumstances^26^. On the other hand, as reported in this study, there were reports of rushed caesarean sections to shorten women’s stay in labour and childbirth. The increased level of medically unjustified caesarean sections during the COVID-19 pandemic might have its own downside. Women and their newborns might have an extended stay in health facilities until they recover which in turn poses a counterproductive effect of substantially increasing their risk of exposure to SARS-CoV-2^26^. Moreover, conducting avoidable caesarean sections uses more of the already limited PPE in health facilities, especially in LMIC settings^17^.

Perceived somewhat or substantially lower ability to provide RMC during the COVID-19 pandemic was higher among high-income country participants and midwives. This could be due to the reality that, in most high-resource settings, pre-existing routine systems or models of care embraced the various dimensions of RMC (companionship, equitable access, evidence-based interventions, etc.); hence, disruptions to existing protocols were noticeable to health workers and reported. The higher levels of RMC disruption reported by midwives might also be due to their salient role in providing maternity care that ensures a balance between women’s rights and experience of care, and the need to prevent infection in health facilities^36^. Midwives’ model and approach to providing continuous emotional support, and informed choice and avoiding unnecessary interventions during pregnancy, labour, childbirth, and the postpartum period might also make it more likely that they noticed and reported lower levels of RMC in their settings. Findings of our first round global survey also strengthen this argument as midwives reported serious concerns about protecting women and newborns^12^. In a study of midwives’ experiences of providing maternity care during the COVID-19 pandemic in Australia, researchers found that stress affected midwives’ ability to provide quality maternity care; midwives experienced direct and indirect stress due to fear of getting infected and infecting others and the potential negative consequences of reduced face to face care on the health of women and newborns^37^. The midwives’ concern is supported by a recent meta-analysis of 40 studies that found a significant increase in maternal death, postnatal depression, and still birth during the COVID-19 pandemic that are partly attributable to delayed and substandard care^38^.

### Implications for health systems

Data were collected starting in July 2020, when basic knowledge was available about transmission of SARS-CoV-2, infection prevention in health facilities, including the use of PPE, very low risk of vertical transmission, and the benefits of breastfeeding and rooming-in for mothers with suspected or confirmed COVID-19^39^. Notwithstanding this knowledge, this study indicates that RMC continued to be critically affected; meaning that the impacts of COVID-19 on RMC might not temporary and require a concerted effort to reverse now and in the future.

Members of the Global RMC Council—a global network of more than 350 experts from 45 countries—also reported wide-ranging violations of women’s rights to RMC globally, including mother-newborn separation and unnecessary caesarean sections, and reiterated the need to strengthen the quality of maternal and newborn health services to protect the rights of women and their newborns^20^. Compromised standards of care, including RMC—as a result of measures taken to limit SARS-CoV-2 transmission especially during the early pandemic when little evidence existed^35^—undermine the trust communities have in the health system and the progress to date to achieve the maternal health targets of the SDGs^13, 38^. Moreover, if prompt mitigative actions are not taken to improve RMC and protect and promote the rights of women in the face of the COVID-19 pandemic, current global inequities in maternal health might be exacerbated^4^. Accordingly, it is imperative to work towards strengthening a health system that is capable of absorbing shocks and preserving the quality of care in times of crises such as the COVID-19 pandemic^4, 10^.

The Ebola virus epidemic in West Africa also devastated the provision of maternal and newborn health services and increased maternal and newborn mortality rates^28, 29^. Additionally, the epidemic resulted in lowering the uptake of facility-based childbirth due to various factors including women experiencing and perceiving disrespectful care and harm related to health workers wearing PPE during maternity care in health facilities^30, 31^. Strengthening public information about patient experience during the Ebola virus epidemic and research on RMC were missed opportunities that could have generated mitigative actions that could be translated to the current pandemic situations^32, 33^.

The entangled nature of the negative impacts of the COVID-19 pandemic on the provision of RMC indicates that these impacts should be addressed from a multidimensional health system strengthening perspective to ensure the sustainability of results^10^. Building local capacity to translate emerging evidence and global guidelines into context-adapted strategies is vital to ensure that essential services will be provided with little or no disruption of RMC^40^. The growing focus on maternity care and opportunities that come along the COVID-19 pandemic, if successfully acted upon, could present innovative and creative approaches to advancing the quality of maternity care generally and RMC particularly, even after the pandemic^34^.

### Strengths and limitations

This study is among the few studies that examined how the COVID-19 pandemic negatively impacted RMC globally from its onset. Our study provides a nuanced understanding of how the COVID-19 pandemic affected RMC across different settings and the potential areas of action that could be focused on to promote RMC and protect the rights of women and newborns in the remaining trajectory of the pandemic—the lessons learned could also be applied to future pandemics and health system disruptions. In our study, nearly half of the participants reported that their ability to provide respectful care was somewhat or substantially improved during the COVID-19 pandemic compared to before. This could be due to various factors including: 1) health workers perceiving certain RMC practices, such as companionship, as not essential to quality of care or not possible during a pandemic, 2) infection prevention measures taken during the pandemic, such as partitioning of rooms, providing more privacy, 3) resources, opportunities, and hygiene measures that evolved during the pandemic and improved the maternal health care system^34^, 4) reduced patient volumes in some settings due to women’s fear of viral infection^35^, resulting in more time and resources per client, and 5) new global guidelines that were introduced to protect the rights of childbearing women during the COVID-19 pandemic.

However, our study is limited in terms of exploring how the ability of health workers to provide RMC improved during the COVID-19 pandemic compared with before the pandemic. This is because the participants who reported their ability to provide RMC during the pandemic was somewhat better or substantially better than before the pandemic were not asked a subsequent open-ended question. Additionally, as the majority (88%) of the participants who responded to the question that inquired about how the COVID-19 pandemic negatively affected RMC were from high-and middle-income countries, the study might be limited in depicting a clear picture of how the pandemic affected RMC in low-income countries. We imagine that in low-resource settings where a focus on RMC was relatively recent, health providers would have experienced quite different impacts; birth companions were not allowed in many settings already, for example. The pre-pandemic models of care in these settings were likely to have been set up to prioritize the concerns and approaches of the health system and health workers, not client experience—so, what happened as a result of the pandemic was not so much of a disruption to how maternity care was provided.

Lastly, our study did not aim to gather women’s or families’ experiences of RMC during the COVID-19 pandemic as all our participants were health workers. Future studies should approach this particular research area using mixed methods design and use these together with the reports of health workers to adequately explore the barriers and enablers to the provision of RMC, included supports to be provided to health workers, during the pandemic and beyond.

## Conclusion

The COVID-19 pandemic adversely affected RMC in a multitude of ways in different settings globally. According to health workers who provide maternal and newborn care, it compromised mainly the clinical and interpersonal elements of care. Women’s right to choose their preferred mode of childbirth, and benefit from companionship during labour, childbirth and postpartum were also affected. Additionally, the pandemic led to fear, confusion, and frustration among health workers limiting their ability to provide RMC according to evidence-based guidelines. These negative impacts were entangled and therefore necessitate multidimensional and system-oriented intervention approaches if women’s rights to RMC are to be protected during the COVID-19 pandemic and thereafter. Furthermore, opportunities that emerge in the era of the COVID-19 pandemic should be capitalised on to foster innovations and interventions that could promote RMC globally.

## Data Availability

Anonymised data analysed during the current study will be made available from the corresponding author upon reasonable request

## Acknowledgement

The authors would like to thank all health workers who participated in this study for giving us time to share their experiences of maternal and newborn care provision during the COVID-19 pandemic. We are also grateful to all individuals and organisations who helped us to promote the global survey to reach health workers across the globe.

## Ethical statement

This study received ethical clearance from the Institutional Review Board at the Institute of Tropical Medicine (approval number: 1372/2020). Participants were provided with information about the study and they gave online written consent before participating in the study. Data were collected anonymously and no personal identifiers were used in reporting findings from the study.

## Author contributions

**Anteneh Asefa:** Conceptualisation, Data curation, Formal analysis, Methodology, Visualisation, Writing – original draft. **Aline Semaan:** Conceptualisation, Data curation, Writing – review & editing. **Therese Delvaux:** Conceptualisation, Writing – review & editing. **Elise Huysmans**: Conceptualisation, Writing – review & editing. **Anna Galle**: Conceptualisation, Writing – review & editing. **Emma Sacks:** Conceptualisation, Formal analysis, Writing – review & editing. **Meghan A. Bohren:** Conceptualisation, Writing – review & editing. **Alison Morgan:** Conceptualisation, Writing – review & editing. **Michelle Sadler:** Conceptualisation, Writing – review & editing. **Saraswathi Vedam:** Conceptualisation, Writing – review & editing. **Lenka Benova:** Conceptualisation, Data curation, Formal analysis, Funding acquisition, Investigation, Methodology, project administration, Resources, Writing – review and editing.

---

**Supplementary file 1.** Distribution of participants who responded to the open-ended respectful care question by country.

| **No.** | **Country** | **Number** | **Percentage** |
| --- | --- | --- | --- |
| 1 | Argentina | 2 | 1.7 |
| 2 | Belgium | 2 | 1.7 |
| 3 | Benin | 1 | 0.8 |
| 4 | Bolivia | 1 | 0.8 |
| 5 | Brazil | 2 | 1.7 |
| 6 | Cameroon | 4 | 3.3 |
| 7 | Canada | 3 | 2.5 |
| 8 | Congo, Democratic Republic of the | 8 | 6.7 |
| 9 | France | 4 | 3.3 |
| 10 | Germany | 11 | 9.2 |
| 11 | India | 6 | 5.0 |
| 12 | Italy | 6 | 5.0 |
| 13 | Japan | 9 | 7.5 |
| 14 | Kazakhstan | 12 | 10.0 |
| 15 | Malawi | 1 | 0.8 |
| 16 | Morocco | 2 | 1.7 |
| 17 | Mozambique | 1 | 0.8 |
| 18 | Nigeria | 8 | 6.7 |
| 19 | Norway | 6 | 5.0 |
| 20 | Panama | 1 | 0.8 |
| 21 | Peru | 1 | 0.8 |
| 22 | Rwanda | 2 | 1.7 |
| 23 | Saudi Arabia | 1 | 0.8 |
| 24 | Spain | 5 | 4.2 |
| 25 | Sweden | 1 | 0.8 |
| 26 | Switzerland | 2 | 1.7 |
| 27 | Tanzania | 3 | 2.5 |
| 28 | Uganda | 2 | 1.7 |
| 29 | United Kingdom | 2 | 1.7 |
| 30 | United States | 5 | 4.2 |
| 31 | Uruguay | 4 | 3.3 |
| 32 | Uzbekistan | 1 | 0.8 |
| 33 | Zambia | 1 | 0.8 |
|  | **Total** | **120** | **100** |

